# The relationship between the size and asymmetry of the lateral ventricles and cortical myelin content in individuals with mood disorders

**DOI:** 10.1101/2024.04.30.24306621

**Authors:** A Manelis, H Hu, R Miceli, S Satz, R Lau, S Iyengar, HA Swartz

## Abstract

**Background:** Although enlargement of the lateral ventricles was previously observed in individuals with mood disorders, the link between ventricular size and asymmetry with other indices of brain structure remains underexplored. In this study, we examined the association of lateral ventricular size and asymmetry with cortical myelin content in individuals with bipolar (BD) and depressive (DD) disorders compared to healthy controls (HC).

**Methods:** Magnetic resonance imaging (MRI) was used to obtain T1w and T2w images from 149 individuals (age=27.7 (SD=6.1) years, 78% female, BD=38, DD=57, HC=54). Cortical myelin content was calculated using the T1w/T2w ratio. Elastic net regularized regression identified brain regions whose myelin content was associated with ventricular size and asymmetry. A post-hoc linear regression examined how participants’ diagnosis, illness duration, and current level of depression moderated the relationship between the size and asymmetry of the lateral ventricles and levels of cortical myelin in the selected brain regions.

**Results:** Individuals with mood disorders had larger lateral ventricles than HC. Larger ventricles and lower asymmetry were observed in individuals with BD who had longer lifetime illness duration and more severe current depressive symptoms. A greater left asymmetry was observed in participants with DD than in those with BD (p<0.01). Elastic net revealed that both ventricular enlargement and asymmetry were associated with altered myelin content in cingulate, frontal, and sensorimotor cortices. In BD, but not other groups, ventricular enlargement was related to altered myelin content in the right insular regions.

**Conclusions:** Lateral ventricular enlargement and asymmetry are linked to myelin content imbalance, thus, potentially leading to emotional and cognitive dysfunction in mood disorders.

## 1 INTRODUCTION

Mood disorders such as depressive (DD) and bipolar (BD) disorders affect millions of people worldwide (Dilsaver, 2011; Greenberg et al., 2021; Kieling et al., 2024). Understanding the neurobiological correlates of these disorders may help improve clinical outcomes. One of the structures affected in individuals with mood disorders is the lateral ventricles (Abé et al., 2023; Gray et al., 2020; Hibar et al., 2016, 2018; Ho et al., 2022; Okada et al., 2023; Schmaal et al., 2016). The lateral ventricles are large C-shaped structures that project into the frontal, temporal, and occipital lobes and are responsible for cerebrospinal fluid (CSF) production (Scelsi et al., 2020). The size of the ventricles positively correlates with the size of the choroid plexus (Murck et al., 2024), which produces CSF and contributes to the maintenance of CNS homeostasis by controlling immune cells and molecule exchange between the CSF and the bloodstream (Thompson et al., 2022).

Although brain anatomical variations are normal in healthy individuals (Scelsi et al., 2020), enlargement of lateral ventricles and ventricular asymmetry may affect the size and structure of the other brain regions located proximally and distally to the lateral ventricles. For example, ventricular enlargement that restricted intracranial space was associated with occipital bending (Kempton et al., 2011), which was higher in individuals with DD (Maller et al., 2014) and those with BD (Maller et al., 2015) compared to HC. Ventricular enlargement was also related to a size reduction in most subcortical regions (Abé et al., 2022), cortical thinning (Chung et al., 2017; Kang et al., 2018), and choroid plexus enlargement (Lizano et al., 2019; Tadayon et al., 2020). The changes in the volume and permeability of the choroid plexus have been associated with neuroinflammation, which, in turn, was linked to altered demyelination and remyelination, especially in the periventricular areas, and the severity of cognitive impairment (Althubaity et al., 2022; Fleischer et al., 2021; Klistorner et al., 2022; Ricigliano et al., 2021; Tadayon et al., 2020; Tonietto et al., 2023).

Previous research proposed that neuroinflammation could be among factors affecting ventricular and choroid plexus enlargement, and mood disorders (Afridi & Suk, 2021; Benedetti et al., 2020; Dantzer et al., 2008; Pereira et al., 2021; Troubat et al., 2021). A recent mega-analysis has shown that individuals with BD and those with DD have significantly larger lateral ventricles compared to HC (Okada et al., 2023). Individuals with BD who had larger lateral ventricles also had a higher number of prior manic episodes (Strakowski et al., 2002). In addition, individuals with BD showed a greater rate of increase in the size of the ventricles over time compared to HC (Abé et al., 2022). Alterations in the choroid plexus were associated with depression (Althubaity et al., 2022; Devorak et al., 2015; Lizano et al., 2019; Turner et al., 2014) and alterations in ventricular size were related to treatment response in DD: non-responders had larger lateral ventricles than responders (Murck et al., 2024).

Brain asymmetry refers to the difference between the size of homologous structures in the left and right hemispheres. Although greater asymmetry of the lateral ventricles was negatively correlated with schizophrenia age of onset (Aso et al., 1995), the studies of brain asymmetry in mood disorders showed inconsistent results. For example, some studies revealed greater cortical and subcortical asymmetry in DD compared to HC (Zuo et al., 2019). However, the recent mega-analysis did not reveal significant group differences in the laterality index of the lateral ventricles in either BD or DD relative to HC (Okada et al., 2023). Considering that emotion processing and regulation were linked to hemispheric asymmetries (Bourne, 2010; Davidson, 1993; Davidson & Irwin, 1999; Turnbull & Salas, 2021) and that both are affected in mood disorders (Joormann & Gotlib, 2010; Miola et al., 2022), understanding ventricular asymmetry may shed light on neurobiological mechanisms underlying emotion dysregulation in BD and DD.

Although most of the studies on neuroinflammation and de/remyelination were conducted in individuals with multiple sclerosis, we believe that this neurobiological mechanism could be generalized to other disorders. No study so far investigated how ventricular enlargement may be associated with the change in the level of cortical myelin in individuals with BD and those with DD. Studies of postmortem samples discovered altered myelin content in individuals affected by these disorders (Simons & Nave, 2016; Valdés-Tovar et al., 2022).

Several in vivo studies reported that individuals with mood disorders have lower levels of myelin compared to HC (Sacchet & Gotlib, 2017; Shim et al., 2023; Zhang et al., 2009). However, our recent work revealed that the altered myelin profile is associated with abnormal reductions of cortical myelin levels in some cortical regions and abnormal elevations in other areas (Baranger et al., 2021a, 2021b).

Although the mechanism by which altered ventricular size drives illness severity in mood disorders remains unclear, one possible explanation is that the changes in ventricular size and asymmetry are associated with the changes in cortical myelin content in brain regions located proximally and distally to the lateral ventricles, thus leading to alterations in neural conduction in the brain. The first goal of this study was to compare the measures of ventricular size and asymmetry in individuals with BD vs. those with DD vs. healthy controls (HC). The second goal was to examine the association between the size and asymmetry of the lateral ventricles and the levels of cortical myelin computed based on the T1/T2 ratio (Glasser et al., 2016). To perform variable (i.e., brain parcels) selection, we used elastic net regularized regression (Friedman et al., 2010; Zou & Hastie, 2005) that was successfully used in our previous studies of mood disorders (Acuff et al., 2019; Baranger et al., 2021a, 2021b; Manelis et al., 2020). This approach combines lasso and ridge regression by removing variables with excessively small coefficients and penalizing excessively large coefficients (Zou & Hastie, 2005), thus allowing to examine models in which the number of explanatory variables (e.g., the number of brain parcels) exceeds the number of samples in the dataset (e.g., the number of participants). We hypothesized that individuals with DD and BD will have larger lateral ventricles than HC and that larger lateral ventricles will be associated with the altered myelin content in the regions proximal to lateral ventricles. Our exploratory analyses examined the relationship between the size and asymmetry of the lateral ventricles, lifetime BD or DD illness duration, medication load, and current depression severity in individuals with mood disorders.

## 2 METHODS

### 2.1 Participants

The study was approved by the University of Pittsburgh Institutional Review Board (IRB number: STUDY20060265). We recruited participants from the community, universities, and counseling and medical centers. All participants were right-handed and fluent in English. They gave written informed consent prior to participating in the study. Patient and HC were matched on age and sex. Individuals with BD and DD met DSM-5 criteria for bipolar disorder (Type I and Type II) and depressive disorders (major depressive (MDD) or persistent depressive (PDD) disorders) accordingly. Healthy controls (HC) had no personal or family history of psychiatric disorders. Exclusion criteria included a history of head injury, metal in the body, pregnancy, claustrophobia, neurodevelopmental disorders, systemic medical illness, premorbid IQ<85 per the National Adult Reading Test (Nelson, 1982), current alcohol/drug abuse (within the previous 3 months), Young Mania Rating Scale scores>10 (YMRS(Young et al., 1978)) at scan, or meeting criteria for any psychotic-spectrum disorder. Participants were excluded from analyses due to diagnosis conversion during the course of the study (n=1 converted to MDD from HC, n=2 converted to BD Type II from MDD), or poor image quality due to motion or issues with the scanner (n=18). Of 38 individuals with BD, 11 were diagnosed with BD Type-I (BD-I) and 27 with BD Type-II (BD-II). Of 57 individuals diagnosed with DD, 38 were diagnosed with major depressive disorder (MDD) and 19 were diagnosed with persistent depressive disorder (PDD).

### 2.2. Clinical assessments

A trained clinician evaluated all participants according to DSM-5 criteria using SCID-5 (First, 2015). The diagnoses were confirmed by a psychiatrist. We also collected the information about medications, lifetime illness onset and duration, number of mood episodes, comorbid psychiatric disorders, current depression symptoms using the Hamilton Rating Scale for Depression (HDRS-25) (Hamilton, 1960), current mania symptoms using the Young Mania Rating Scale (YMRS) (Young et al., 1978), and lifetime depression and hypo/mania spectrum symptomatology using the Mood Spectrum Self Report (MOODS-SR) (Dell’Osso et al., 2002). For each participant, we calculated a total psychotropic medication load (Hassel et al., 2008; Manelis et al., 2016). Greater number of medication and higher doses corresponded to a greater medication load.

### 2.3 Neuroimaging data acquisition

The neuroimaging data were collected at the University of Pittsburgh/UPMC Magnetic Resonance Research Center using a 3T Siemens Prisma scanner with a 64-channel receiver head coil. The DICOM image files were named according to the ReproIn convention (Visconti di Oleggio Castello et al., 2020) and were converted to BIDS dataset using *heudiconv* (Halchenko et al., 2019) and *dcm2niix* (Li et al., 2016). High-resolution T1w images were collected using the MPRAGE sequence with TR=2400ms, resolution=0.8x0.8x0.8mm, 208 slices, FOV=256, TE=2.22ms, flip angle=8°. High-resolution T2w images were collected using TR=3200ms, resolution=0.8x0.8x0.8mm, 208 slices, FOV=256, TE=563ms. Field maps were collected in the AP and PA directions using the spin echo sequence (TR=8000, resolution=2x2x2mm, FOV=210, TE=66ms, flip angle=90°, 72 slices).

### 2.4 Data analyses

#### 2.4.1 Clinical data analysis

Demographic and clinical variables for BD, DD, and HC were compared using ANOVA, t-tests, and chi-square tests (whichever was appropriate) in R (https://www.r-project.org/).

#### 2.4.2 Neuroimaging data processing

Structural data were analyzed using Freesurfer v7.4.0 (Dale et al., 1999). Cortical myelin was calculated based on the T1w/T2w ratio using the minimal preprocessing pipelines developed by the Human Connectome Project (Glasser et al., 2013, 2016; Glasser & van Essen, 2011) with w*orkbench v1.4.2* and *HCPpipelines-4.1.3* installed on a workstation with GNU/Linux Debian 10 operating system. Bias field correction was performed using a pair of spin echo field maps in *PreFreeSurfer*. The images were registered to standard space using MSMSulc (Robinson et al., 2018) in *PostFreeSurfer*. The 360 region Glasser Atlas (Glasser et al., 2016) was used to parcellate the resulting cortical maps.

Quality of T1w and T2w images was visually inspected and examined using *mriqc version 0.15.1* (Esteban et al., 2017). A total of 21 participants were removed from the structural data analyses due to the poor quality of T1w images. Additionally, 8 participants were removed due to poor quality of T2 images or resulting cortical myelin maps. Due to known issues with acquisition quality in brain regions susceptible to artifacts, we identified outlier parcels based on the analysis of variance in each of 360 parcels using Rosner test (“*rosnerTest”* function in R) for multiple outliers (Rosner, 1983).

#### 2.4.3 Neuroimaging data analysis

##### 2.4.3.1 Comparing the lateral ventricles and laterality index among individuals with BD, DD and HC

The volume of the right and left lateral ventricles and choroid plexus were adjusted to the estimated total intracranial volume (ventricular volume/ estimated total intracranial volume)*100. The laterality index was calculated as (L-R)/(L+R) (Okada et al., 2023; Seghier, 2008). A mixed effects model predicted the lateral ventricle volume as a function of the hemisphere (right/left) and the diagnostic group (BD/DD/HC) with age, sex and IQ as covariates and participants as a random factor.

##### 2.4.3.2 Elastic net regression

Elastic net regularized regression (Friedman et al., 2010; Zou & Hastie, 2005) with α=0.5 was conducted across all individuals and used to identify brain regions (i.e., parcels) whose cortical myelin content was associated with the lateral ventricles size or asymmetry. For the analysis of the ventricular size, the right and the left lateral ventricles were combined. Nested cross-validation allowed to avoid model overfitting and bias (Baranger et al., 2021a, 2021b). In each loop of nested cross-validation, one participant of 141 was held out and the the rest of 140 participants were used in the elastic net regression with leave-one-out cross-validation thus resulting in 141 sparse models describing the relationship between cortical myelin and the lateral ventricles size and asymmetry. In order to identify feature stability, we calculated the percent of models that selected a particular brain parcel. For example, if a brain region “A” was selected in all 141 models, the feature stability for the region “A” is 100%. In contrast, if a region “B” was selected in only one of 141 models, the feature stability for the region “B” is 0.7%.

In our previous study, we used label permutation to identify the cutoff value for feature stability. The frequency of label selection for randomly permuted labels was 3.8% (Baranger et al., 2021a, 2021b). In the current study, we decided to go with a more stringent approach and only consider the regions that were selected by more than 10% of models. In the post-hoc analysis, we used all parcels selected by more than 10% of nested elastic net regressions as predictors in the linear regression analysis (separately for ventricular size and asymmetry). While we do not know whether the myelin content affects the ventricular size and asymmetry or the other way around, we further explored the effect of the diagnostic group and the measure of the lateral ventricles on cortical myelin content (i.e., myelin∼Group*ventricular size or asymmetry).

##### 2.4.3.3 Exploratory analysis

Exploratory analyses included linear regression to predict the size and the laterality index of the lateral ventricles from the interaction between group, hemisphere, HDRS25, and illness duration in BD and DD. In the separate analyses, we explored the effect of medications, diagnostic subgroups (BD Type-I, BD Type-II, MDD and PDD), and spectral measures of depression and mania on the size and asymmetry of the lateral ventricles.

## 3 RESULTS

### 3.1 Demographics and clinical characteristics

A total of 149 participants were included in the analyses of the lateral ventricles. Detailed demographics and clinical information are reported in Table 1. The three groups did not differ in sex composition and had marginally significant differences in IQ with individuals with BD having marginally higher IQ than HC. There was a significant difference in mean age among the groups: individuals with BD were significantly younger than HC (p=0.01). As expected, both patient groups had a higher burden of current and lifetime symptoms of depression and mania, compared to HC (p<0.001). At the time of the scan, individuals with DD were marginally more depressed than those with BD (p=0.05) but did not differ from them in the level of mania symptoms. The patient groups did not differ from each other in age of illness onset and illness duration as well as the number of participants with comorbid psychiatric disorders. More individuals with BD were taking psychotropic medications compared to those with DD (p=0.007). Among those who were on psychotropic medications, individuals with BD had a higher medications load (i.e., medload) than individuals with DD (p<0.001). Individuals with BD also had significantly more severe spectral symptoms of depression, mania, and psychopathology in general than those with DD (all p values<0.05).

**Table 1.**
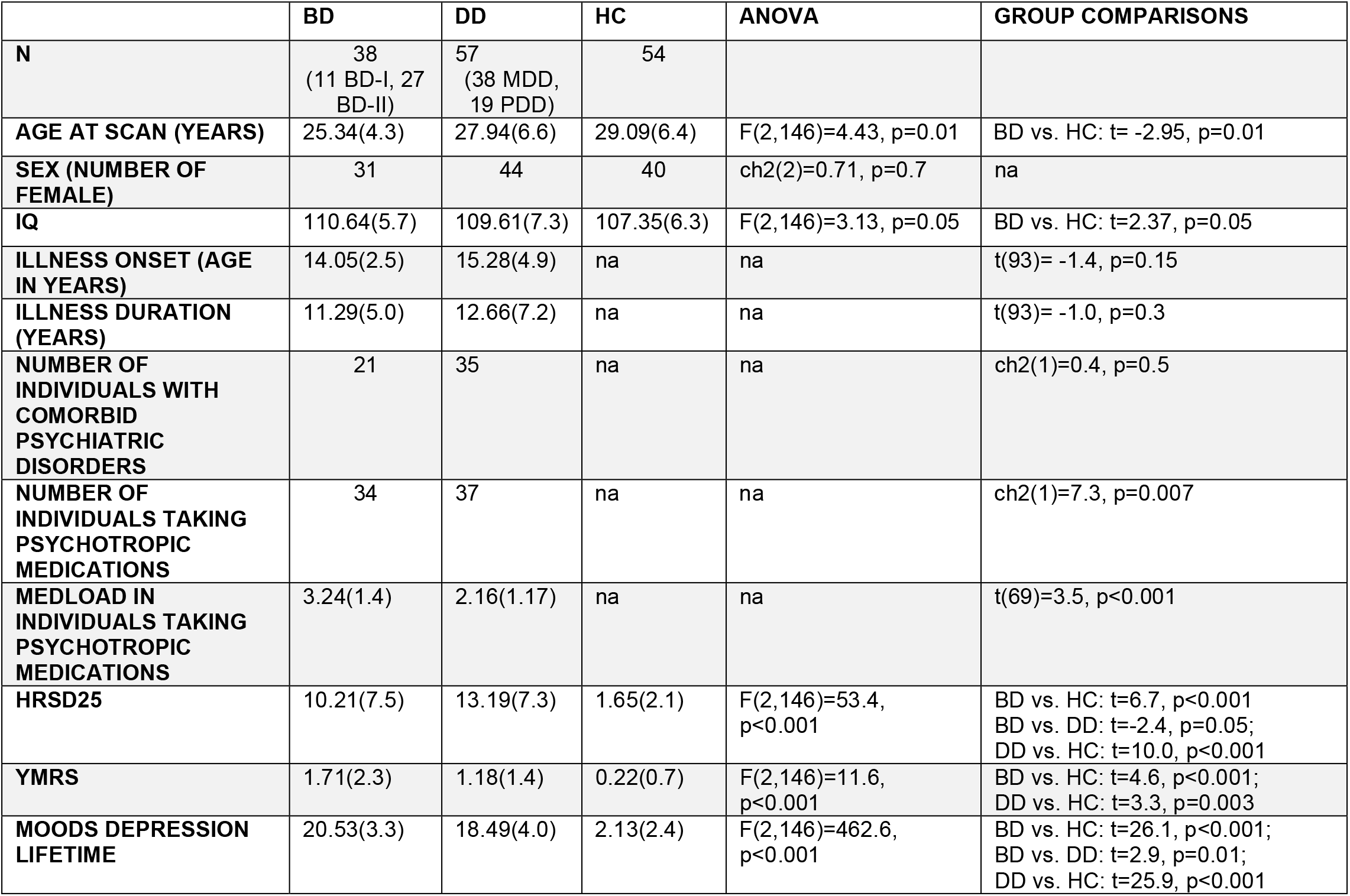
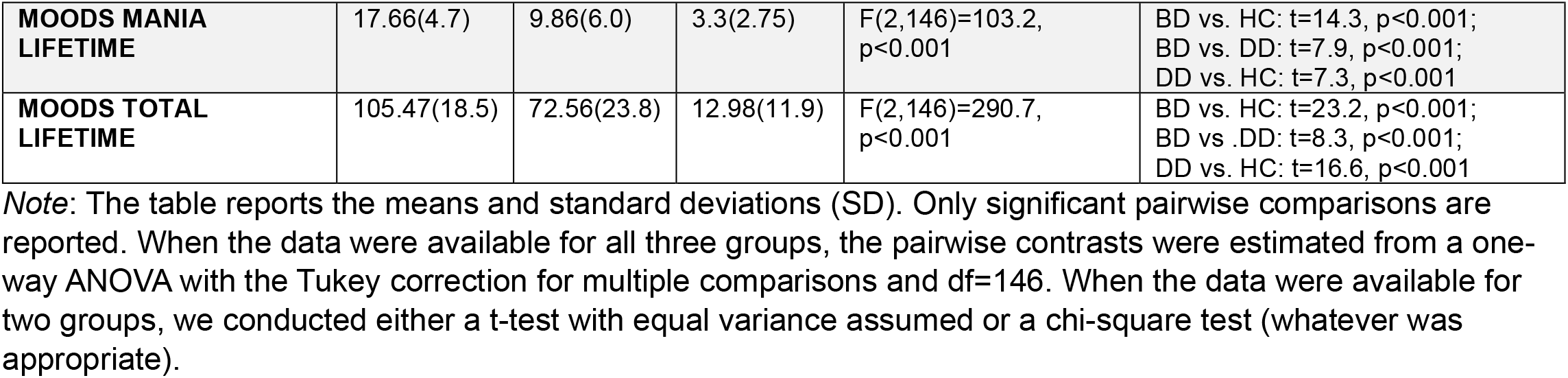

### 3.2 Volume of lateral ventricles and laterality index

There was a significant main effect of group (F(2,143)=4.1, p=0.018) and hemisphere (F(1,146)=14.3, p<0.001) on the adjusted volume of lateral ventricles (Figure 1A). HC had smaller ventricles than BD (t= -2.43, Tukey-corrected-p=0.04) and DD (t= -2.53, Tukey-corrected-p=0.03) who did not differ from each other. Across all participants, the left lateral ventricle was larger than the right lateral ventricle (t=3.8, Tukey-corrected-p<0.001). There was a marginally significant group-by-hemisphere interaction effect (F(2,146)=2.5, p=0.08). There was no significant effect of age, sex, or IQ on the size of the lateral ventricles (all p>0.1).

**Figure 1.**
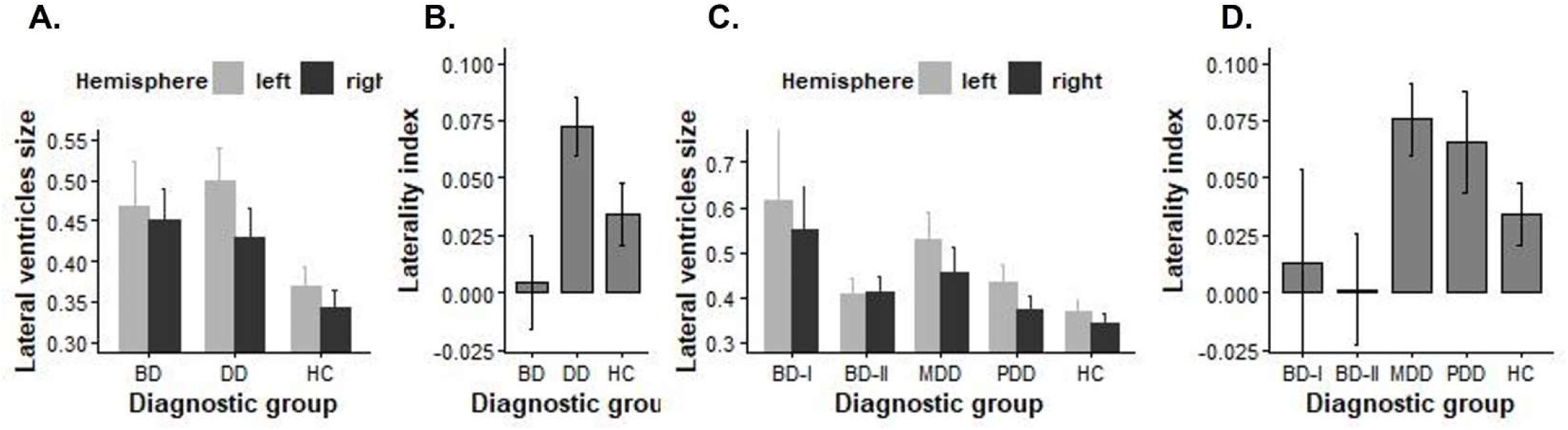
Size and asymmetry of the lateral ventricles in the BD, DD and HC groups. The plots A. and C. depict the mean volume of the right and left lateral ventricles adjusted for the estimated total intracranial volume. The plots B. and D. depict the mean laterality index in each diagnostic group. The error bars are the standard errors of mean. BD – individuals with bipolar disorder, DD – individuals with depressive disorders, HC – healthy controls.

A positive correlation was observed between the sizes of lateral ventricles and choroid plexus (r=0.65, p<0.001). There were significant main effects of group (F(2,143)= 4.2, p= 0.02) and hemisphere (F(1,146)= 23.7, p<0.001) as well as the hemisphere-by-group interaction (F(2,146)= 3.3, p= 0.04) on the choroid plexus size. HC had smaller choroid plexus than individuals with DD (t= -2.7, Tukey-corrected-p=0.02). There was a significant effect of age (F(1,143)=7.4, p=0.007), but no effect of IQ or sex, on the size of choroid plexus.

There was a significant effect of group on the laterality index (F(2,143)=4.8, p<0.01; Figure 1B) with a significantly higher index observed in DD group, compared to BD group (t=3, Tukey-corrected-p=0.009). To better understand the lateral ventricles asymmetry in each group, we compared the laterality index in each group against zero. We found that in the BD group, the laterality index did not differ from the zero (t(37)=0.23, p>0.5) suggesting no asymmetry in the lateral ventricles. However, the laterality index was significantly above zero in both HC (t(53)=2.53, p=0.014) and DD (t(56)=5.7, p<0.001) groups thus suggesting that the left ventricle is larger than the right ventricle.

### 3.3 Exploratory analyses

Exploratory analyses with diagnostic subgroups (BD-I, BD-II, MDD, PDD and HC) showed a significant main effect of group (F(4,141)=3.4, p=0.01) and hemisphere (F(1,144)=15.5, p<0.001) on the adjusted volume of lateral ventricles (Figure 1C). HC had smaller ventricles than individuals with BD-I (t= -3.03, Tukey-corrected-p=0.02) and those with MDD (t= -2.9, Tukey-corrected-p=0.04). The other subgroups did not differ from each other and HC. The analysis of the laterality index for diagnostic subgroups did not yield significant results (Figure 1D).

The rest of the exploratory analyses were conducted in BD and DD groups only to examine the relationship between current depression severity (based on the HDRS25 (Hamilton, 1960) scores) and illness duration on the lateral ventricles size and asymmetry. A mixed effects model for the lateral ventricles size revealed a hemisphere-by-group-by-HDRS25-by-illness duration interaction (F(1,87)=6.8, p=0.01), hemisphere-by-group interaction (F(1,87)=7.4, p=0.008), hemisphere-by-HRSD25 interaction (F(1,87)=8.0, p=0.006), hemisphere-by-illness duration interaction (F(1,87)=6.4, p=0.01), hemisphere-by-Group-by-HRSD25 interaction (F(1,87)=9.6, p=0.003), hemisphere-by-HRSD25-by-illness duration interaction (F(1,87)=4.8, p=0.03) effects (Figure 2). A mixed effects model for the laterality index reveled a significant group-by-HDRS25-by-illness duration interaction effect (F(1,84)=11.9, p<0.001) and a main effect of group (F(1,84)=10.0, p=0.002).

**Figure 2.**
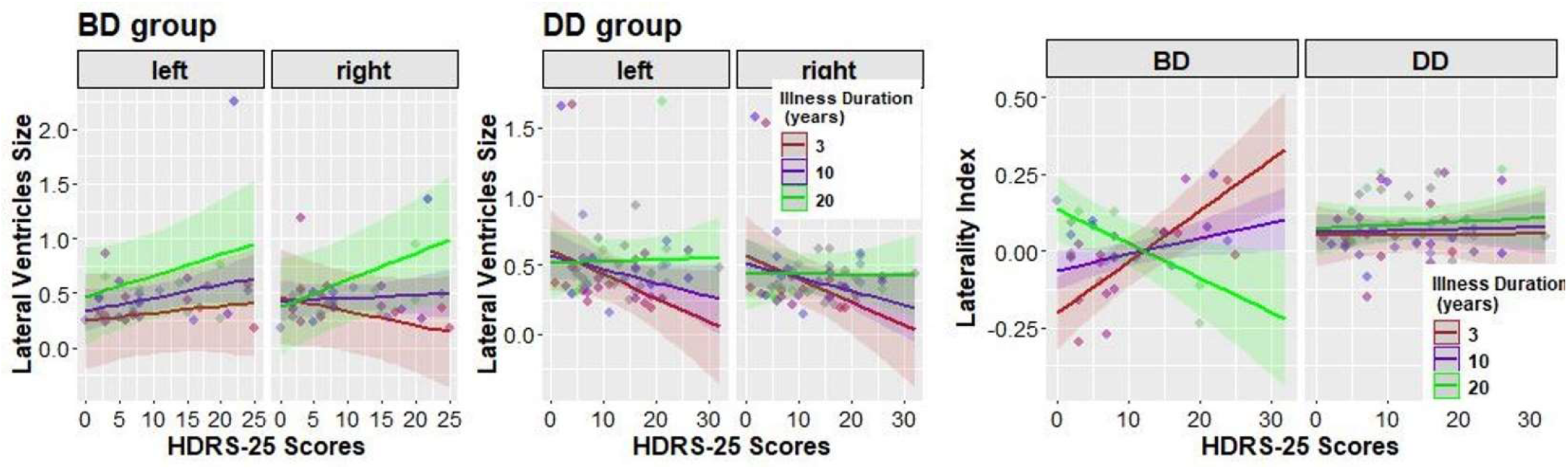
The effect of diagnosis (BD vs. DD), illness duration, and current depression severity based on the HDRS-25 scores on the lateral ventricles size and asymmetry.

The follow-up analyses conducted separately in DD and BD groups revealed no significant main or interaction effects in DD in either lateral ventricles size or asymmetry models. In BD, there was a significant hemisphere-by-HDRS25-by-illness duration interaction effect in the ventricular size model (F(1,34)=4.3, p<0.05). This effect was driven by the right hemisphere that exhibit a significant HRSD25-by-illness duration interaction F(1,31)=4.8, p<0.05). No such interaction or main effects were observed in the left hemisphere. Also, there was a significant HDRS25-by-illness duration interaction effect in the laterality index model (F(1,31)=13.3, p<0.001). A positive association between the laterality index and HDRS25 score was observed in the individuals with BD who had shorter illness duration, but a negative association between these variables was observed in those with BD who had longer illness duration (Figure 2).

There was no main or interaction effect of medication load or taking (vs. not taking) psychotropic medications on the lateral ventricles size or laterality index.

### 3.3 The relationship between the size and asymmetry of lateral ventricles and the level of cortical myelin

Note: All brain parcels described in this section are listed in Table S1 along with their full names and function.

#### 3.3.1 Elastic net regularized regression

Rosner’s outlier test identified 13 outlier parcels whose coefficient of variation significantly differed from the mean coefficient of variation. These regions included R_Pir, R_pOFC, L_Pir, R_52, R_EC, R_PreS, R_47s, L_pOFC, L_PreS, R_H, R_25, L_EC, L_H were excluded from all further analyses of cortical myelin. A total of 141 participants were included in the cortical myelin analysis (36 BD, 54 DD, 51 HC). Eight participants were excluded due to poor quality of T2w images or myelin maps.

##### 3.3.1.1 Size of the lateral ventricles and cortical myelin

Elastic net regularized regression selected 20 brain parcels whose cortical myelin content was associated with the lateral ventricles size (Figure 3A). Each of these parcels was selected by at least 10% of the nested cross-validation models (Table 2). The post-hoc linear regression analysis revealed that these 20 variables explained 62.8% of variance in the size of the Lateral Ventricles (F(20,120)=10.1, p<0.001). The regression coefficients estimated for each predictor in this regression model are reported in Table 2 and Figure 3B. A significant interaction effect of bilateral ventricles size and group on cortical myelin content was found in R_23c, R_AVI, R_Ig, and R_RSC (Table 2, Figure 3C). There was a significant main effect of the lateral ventricles size in most of the selected parcels. A significant group effect was observed only in R_31pd, R_24dv, R_STSdp, and L_11l.

**Table 2.** The relationship between the lateral ventricles size and the level of cortical myelin: the results of regularized regression and post-hoc linear regression analysis

| Parcel | Elastic net<br>Selection<br>frequency<br>% | Full<br>regressi<br>on<br>model<br>Estimat<br>es | Cortical myelin level ~ group * bilateral lateral ventricles size |  |  |
| --- | --- | --- | --- | --- | --- |
|  |  |  | Main effect of lateral<br>ventricles size | Main effect of group | Lateral ventricles size *<br>Group interaction |
| L_24dv | 100 | -3.9* | **F(1,135)=15.16,p<0.001** | F(2,135)=1.58,p=0.2 | F(2,135)=1.99,p=0.1 |
| L_RI | 100 | -2.5* | **F(1,135)=18.12,p=<0.001** | F(2,135)=2.42,p=0.1 | F(2,135)=0.84,p=0.4 |
| R_RSC | 100 | -2.8* | **F(1,135)=25.76,p<0.001** | F(2,135)=0.23,p=0.8 | **F(2,135)=5.64,p=0.004** |
| R_VVC | 100 | 1.9* | **F(1,135)=10.27,p=0.002** | F(2,135)=0.84,p=0.4 | F(2,135)=0.07,p=0.9 |
| L_DVT | 99.3 | -1.9 | **F(1,135)=5.36,p=0.02** | F(2,135)=0.52,p=0.6 | F(2,135)=1.26,p=0.3 |
| L_PSL | 99.3 | 2.9* | **F(1,135)=7.07,p=0.01** | F(2,135)=0.07,p=0.9 | F(2,135)=0.11,p=0.9 |
| L_SCEF | 99.3 | 7.5* | **F(1,135)=10.98,p=0.001** | F(2,135)=0.05,p=0.9 | F(2,135)=2.5,p=0.09 |
| R_31pd | 99.3 | 2.7 | **F(1,135)=10.57,p=0.002** | **F(2,135)=3.68,p=0.02** | F(2,135)=0.73,p=0.5 |
| R_33pr | 99.3 | -1.3 | **F(1,135)=15.26,p<0.001** | F(2,135)=0.15,p=0.9 | F(2,135)=1.48,p=0.2 |
| R_AVI | 99.3 | -2.6* | **F(1,135)=12.79,p<0.001** | F(2,135)=1.22,p=0.3 | **F(2,135)=5.32,p=0.006** |
| R_Ig | 99.3 | 1.6 | **F(1,135)=12.82,p<0.001** | F(2,135)=1.87,p=0.2 | **F(2,135)=12.65,p<0.001** |
| R_p24 | 99.3 | -1.9 | **F(1,135)=10.58,p=0.001** | F(2,135)=0.28,p=0.8 | F(2,135)=1.74,p=0.2 |
| R_1 | 98.6 | -1.6 | **F(1,135)=5.58,p=0.02** | F(2,135)=1.17,p=0.3 | F(2,135)=2.06,p=0.1 |
| R_23c | 98.6 | 2.1 | **F(1,135)=9.82,p=0.002** | F(2,135)=0.05,p=0.95 | **F(2,135)=3.86,p=0.02** |
| R_24dv | 97.9 | 3.9* | F(1,135)=3.87,p=0.05 | **F(2,135)=3.13,p=0.05** | F(2,135)=0.56,p=0.6 |
| R_6r | 97.9 | -5.2* | **F(1,135)=5.13,p=0.03** | F(2,135)=0.92,p=0.4 | F(2,135)=0.38,p=0.7 |
| R_FOP4 | 97.2 | -0.7 | **F(1,135)=9.38,p=0.003** | F(2,135)=0.37,p=0.7 | F(2,135)=0.61,p=0.5 |
| R_STSdp | 85.1 | 1.7 | **F(1,135)=5.73,p=0.02** | **F(2,135)=3.34,p=0.04** | F(2,135)=2.48,p=0.09 |
| L_25 | 59.6 | 0.8 | **F(1,135)=4.17,p=0.04** | F(2,135)=0.01,p=0.99 | F(2,135)=0.61,p=0.5 |
| L_11l | 56.7 | -3.1 | **F(1,135)=5.34,p=0.02** | F(2,135)=3.07,p=0.05 | F(2,135)=1,p=0.4 |

Note: \* - significant at $p<0.05$

**Figure 3.**
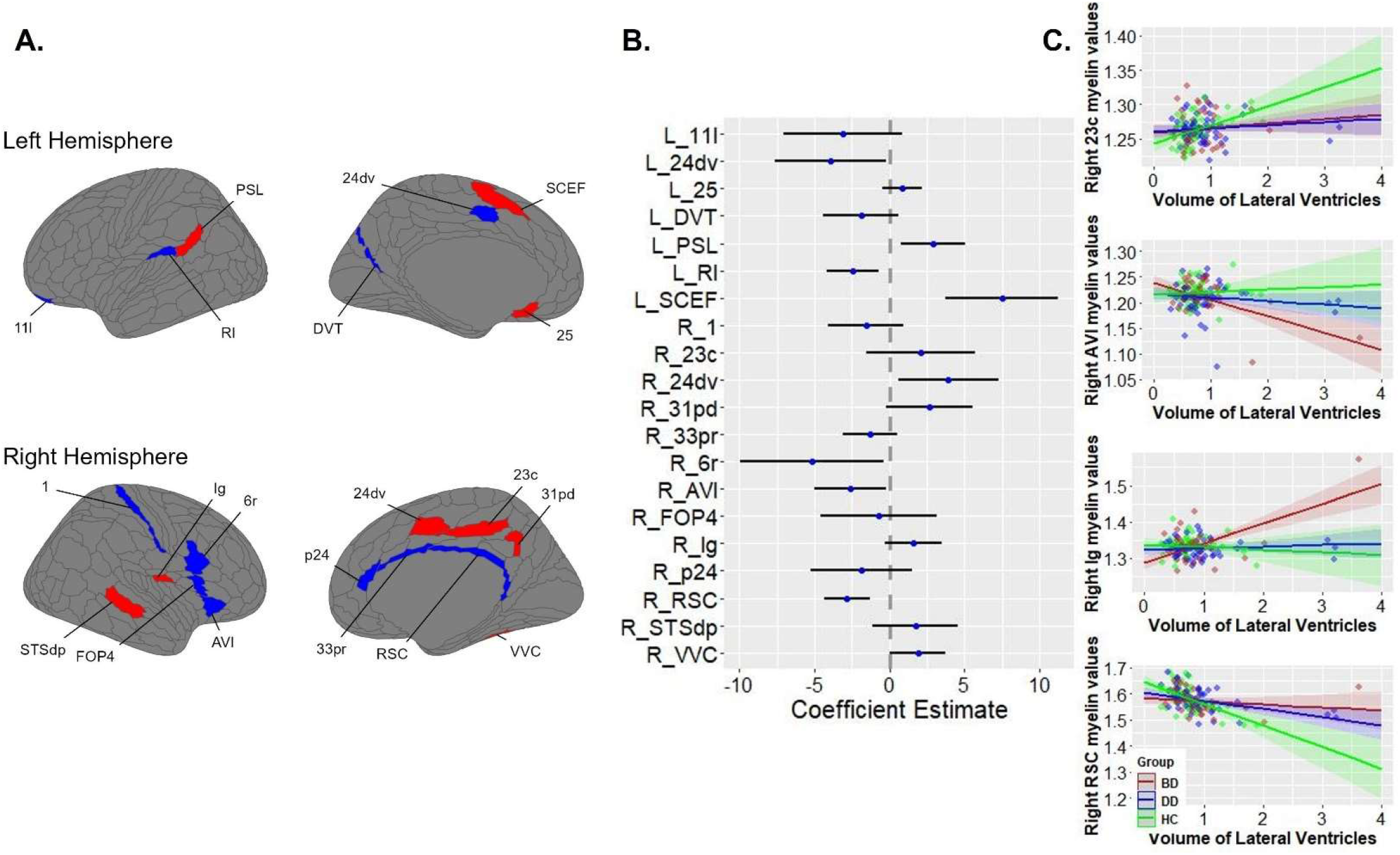
The association between the lateral ventricles size and cortical myelin content. (A) Brain parcels selected by regularized regression. (B) Regression coefficients with 95% confidence intervals for each brain parcel depicted in Figure 2A. (C) The relationship between the lateral ventricles size and cortical myelin content as a function of participants’ diagnostic status. BD – individuals with bipolar disorder, DD – individuals with depressive disorders, HC – healthy controls.

##### 3.3.1.2 Ventricular asymmetry and cortical myelin

Elastic net regularized regression revealed that the laterality index was related to cortical myelin content in 9 parcels (Figure 4A) that were selected as important variables by more than 10% of models (Table 3). The post-hoc linear regression analysis with all 9 parcels revealed that these variables explained 30.3% of variance in the laterality Index (F(9,131)=6.3, p<0.001; Figure 4B). A significant interaction effect of the laterality index and group on myelin levels was found in L_PHA2, R_PHT, R_5mv, and R_OFC (Table 3, Figure 4C). There was a significant main effect of laterality index on myelin levels in all selected parcels. A significant group effect was observed only in R_7Pm.

**Table 3.**
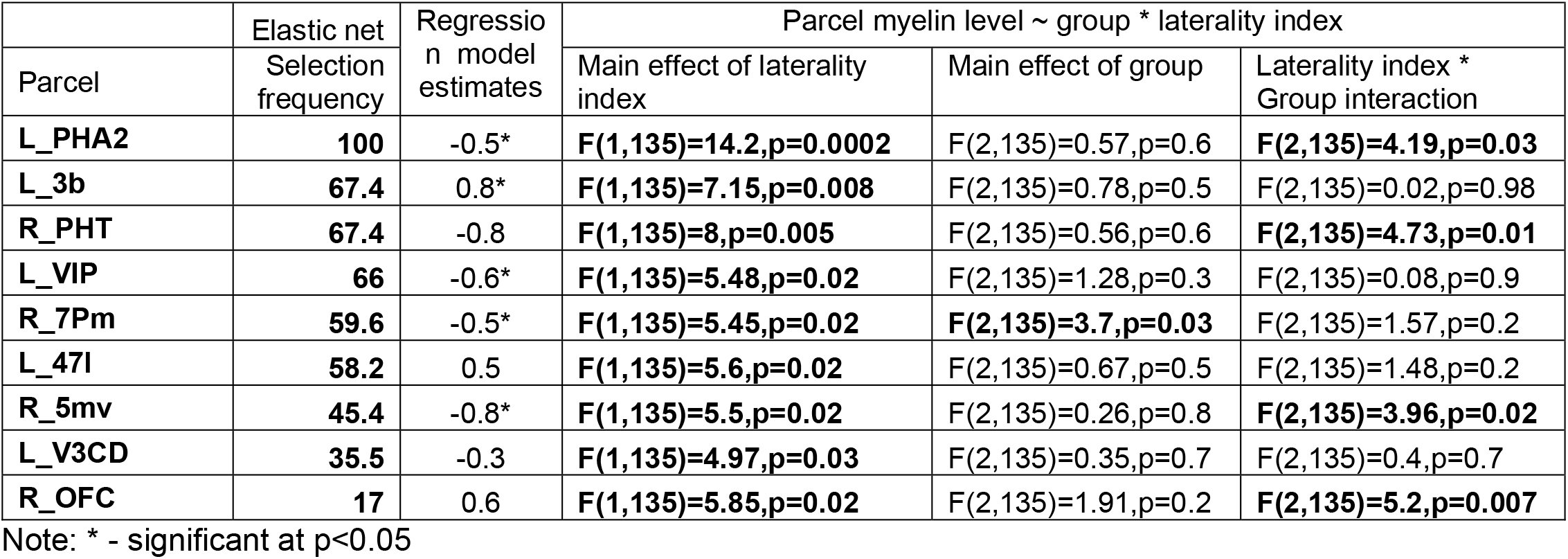

**Figure 4.**
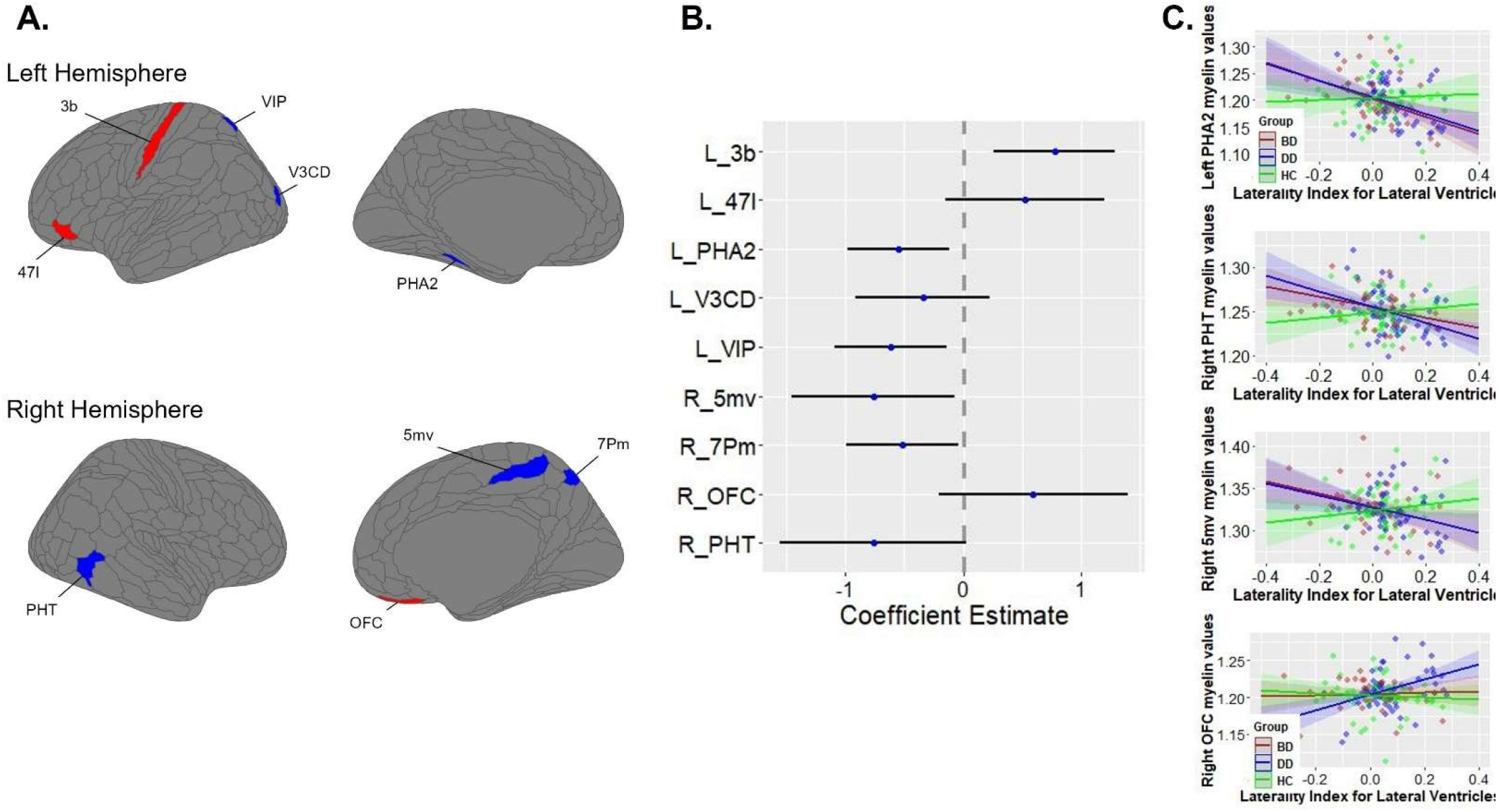
The association between the laterality index and cortical myelin content. (A) Brain parcels selected by regularized regression. (B) Regression coefficients with 95% confidence intervals for each brain parcel depicted in Figure 3A. (C) The relationship between the laterality index and cortical myelin content as a function of participants’ diagnostic status. BD – individuals with bipolar disorder, DD – individuals with depressive disorders, HC – healthy controls.

## DISCUSSION

In this study, we compared the size and asymmetry of the lateral ventricles in individuals with BD and DD relative to each other and HC. We also examined the relationship between the lateral ventricles size and asymmetry with the levels of cortical myelin observed in the brain regions proximal and distal to the lateral ventricles. Consistent with previous studies, the individuals with mood disorders had larger lateral ventricles than their HC counterparts (Hibar et al., 2016; Okada et al., 2023; Schmaal et al., 2016; Strakowski et al., 2002). This distinction was the most pronounced in the individuals with BD-I and MDD relative to HC rather than those with BD-II or PDD, thus reflecting the relationship between ventricular enlargement and mood disorder specificity (Edmiston et al., 2011; Grewal et al., 2023; Hauser et al., 2000; Scott et al., 1983). Our further investigation into the relationship between the lateral ventricles size, illness duration, and current depression severity revealed no relationship among these variables in individuals with DD. In those with BD, the right, but not left, ventricle was larger in currently depressed individuals with longer illness duration relative to those with shorter illness duration. These findings align with those suggesting the relationship between the size of the right lateral ventricle and the number of illness episodes in individuals with BD (Brambilla et al., 2001).

Consistent with previous studies, the laterality index in individuals with mood disorders was not different from that in HC (De Kovel et al., 2019; Okada et al., 2023). A novel finding was that the laterality index distinguished individuals with BD from those with DD. While the left hemisphere dominance characterized individuals with DD, the laterality index in those with BD did not differ from zero, suggesting a lack of asymmetry in the lateral ventricles. Furthermore, the exploratory analysis showed that the laterality index in individuals with BD changed as a function of current depression severity and illness duration. The left asymmetry was greater in depressed individuals with BD and shorter lifetime illness duration than in those with longer illness duration. There was no relationship between the laterality index, illness duration, and depression severity in individuals with DD. We propose that the lack of asymmetry in individuals with BD could be related to the alternations between depression and mania leading to enlargement of the right lateral ventricle, thus decreasing ventricular asymmetry. Future longitudinal studies should examine how brain asymmetry is related to experiencing episodes of mania.

Although the exact causes of ventricular enlargement are still unknown, neuroinflammation was proposed as one of the factors affecting ventricular size and leading to ventricular wall degeneration. Such degeneration may result in diffusion of potentially toxic cerebrospinal fluid through the walls, damage to the periventricular white matter (Coutu et al., 2015; Tonietto et al., 2023; Yamada & Mase, 2023), and disruption of the blood-brain barrier (BBB) (Alber et al., 2019). Additionally, enlargement of the lateral ventricles may exert pressure on periventricular areas, potentially making the adjacent periventricular white and deep grey matter susceptible to compression (Mierzwa et al., 2015; Murck et al., 2023; Sinnecker et al., 2020). It was proposed that both bipolar (Benedetti et al., 2020; Pereira et al., 2021) and depressive (Afridi & Suk, 2021; Dantzer et al., 2008; Troubat et al., 2021) disorders may have the neuroinflammatory basis potentially associated with the immune system response to stress (Glaser & Kiecolt-Glaser, 2005; Slavich & Irwin, 2014) and leading to more severe symptoms of depression (Felger & Miller, 2020). In this study, we observed a strong positive correlation between the sizes of the lateral ventricles and the choroid plexus that were larger in individuals with DD than HC. Previous studies linked the choroid plexus enlargement with disruption of the BBB and depression (Althubaity et al., 2022; Turkheimer et al., 2021), but also with altered demyelination and remyelination processes with more pronounced demyelination occurring in the regions proximal to the ventricles (Tonietto et al., 2023).

In line with these findings, we showed that enlargement of the lateral ventricles was associated with *lower* levels of cortical myelin along the cingulate cortex (R p24, R 33pr, R RSC) *proximal* to the lateral ventricles and adjacent to the periventricular white matter. In addition, the lateral ventricles size was associated with *lower* levels of cortical myelin, in the in the frontal cortical areas responsible for working memory and language (R 6r, R FOP4, L11l), parieto-occipital sulcus (L DVT), primary somatosensory cortex (area 1), and dorsal cingulate area L 24dv. The lateral ventricles size was also associated with *higher* levels of cortical myelin in the dorsal (R 24dv, R 23c, R 31pd) and rostral (L 25) cingulate cortex, insula (R Ig, L PoI1), and the brain regions involved in cognitive function and language (L PSL, L SCEF, and R STSdp). These areas were located more *distally* from the ventricles than areas showing a negative association with myelin levels. Similar to our previous study (Baranger et al., 2021a), some brain regions with higher myelin content were adjacent to those with lower myelin content, thus possibly reflecting a compensatory mechanism.

Interestingly, larger lateral ventricles were related to lower myelin content in the R RSC and higher myelin content in the R 23c (both are the parts of the cingulate cortex) in HC, but not individuals with mood disorder. Larger lateral ventricles were related to lower myelin content in the R AVI and higher myelin content in the R Ig (both are the parts of the insula) in BD but not in HC or individuals with DD, thus suggesting an imbalance between different parts of the right insula in BD that may result in aberrant sensory-motor, emotional, and cognitive processes supported by the insula (Baker et al., 2018). Considering that a reduction in cortical myelin may be associated with decreased connectivity of the affected region and the rest of the brain, our findings may explain previously reported reduced function connectivity between the right AVI and parietal regions in individuals with BD (Qiu et al., 2020).

The relationship between cortical myelin content and brain asymmetry was, in general, consistent with hemisphere dominance. The individuals with higher laterality index and, consequently, larger left vs. right lateral ventricles, showed *higher* myelin content in the left primary sensory (area 3b), language (area 47l), and reward evaluation (R OFC) brain regions than the individuals with lower laterality index. A negative relationship was observed between the laterality index and the regions involved in memory (R PHT, L PHA2), attention (L VIP), and object recognition (L V3CD). Interestingly, despite the significant differences in the laterality index in BD vs. DD, cortical myelin content in the LPHA2, RPHT, and R5mv showed a reduction with the increase in the laterality index in BD and DD, but not HC. These results may explain variability in symptoms of memory dysfunction in individuals with mood disorders: those with greater asymmetry in lateral ventricles, and consequently, lower myelin content in parahippocampal (PHA2) and posterior middle temporal (PHT) cortices, might experience greater memory disturbance that those with lower ventricular asymmetry. Future studies should examine individual differences in memory as a function of ventricular asymmetry and mood disorder diagnosis. The R OFC showed a positive correlation with the laterality index in the individuals with DD, but not in those with BD or HC. Given that the medial OFC is activated by rewarding stimuli (Rolls et al., 2020) and shows reduced reward sensitivity in depressed individuals (Xie et al., 2021), we hypothesize that asymmetry-related increases in R OFC cortical myelin in DD may help increase neural conduction in this region thus improving reward sensitivity and compensating for myelin decreases in the other brain regions.

In summary, the present study lays the foundation for understanding alterations in lateral ventricular size, asymmetry, and myelin content among individuals with BD, DD, and HC. Lateral ventricular size and asymmetry emerge as neurobiological correlates associated with variations in myelin content in general and in mood disorders specifically. Future longitudinal studies should examine the time course of the changes in ventricular size and asymmetry as the disease progresses. They should also prospectively examine how the changes in the severity of current depression (HDRS-25) would be related to the change in the size of the lateral ventricles.

## Data Availability

All data produced in the present study are available upon reasonable request to the authors after the manuscript is published in a peer reviewed journal

## ACKNOWLEDGMENTS

Funding Acknowledgements: This work was supported by a grant from the National Institute of Health R01MH114870 to A.M. The authors declare no conflict of interest.

H.A.S: receives royalties from Wolters Kluwer, royalties and an editorial stipend from APA Press, and honoraria from Webscape/WebMD, Clinical Education Alliance, and Mediflix.

## REFERENCES

Abé, C., Ching, C. R. K., Liberg, B., Lebedev, A. V., Agartz, I., Akudjedu, T. N., Alda, M., Alnæs, D., Alonso-Lana, S., Benedetti, F., Berk, M., Bøen, E., Bonnin, C. del M., Breuer, F., Brosch, K., Brouwer, R. M., Canales-Rodríguez, E. J., Cannon, D. M., Chye, Y., … Landén, M. (2022). Longitudinal Structural Brain Changes in Bipolar Disorder: A Multicenter Neuroimaging Study of 1232 Individuals by the ENIGMA Bipolar Disorder Working Group. Biological Psychiatry, 91(6), 582–592. 10.1016/j.biopsych.2021.09.008

Abé, C., Liberg, B., Klahn, A. L., Petrovic, P., & Landén, M. (2023). Mania-related effects on structural brain changes in bipolar disorder – a narrative review of the evidence. Molecular Psychiatry, 28(7), 2674–2682. 10.1038/s41380-023-02073-4

Acuff, H. E., Versace, A., Bertocci, M. A., Ladouceur, C. D., Hanford, L. C., Manelis, A., Monk, K., Bonar, L., McCaffrey, A., Goldstein, B. I., Goldstein, T. R., Sakolsky, D., Axelson, D., Birmaher, B., & Phillips, M. L. (2019). Baseline and follow-up activity and functional connectivity in reward neural circuitries in offspring at risk for bipolar disorder. Neuropsychopharmacology, 44(9), 1570–1578. 10.1038/s41386-019-0339-2

Afridi, R., & Suk, K. (2021). Neuroinflammatory Basis of Depression: Learning From Experimental Models. In Frontiers in Cellular Neuroscience (Vol. 15). Frontiers Media SA. 10.3389/fncel.2021.691067

Alber, J., Alladi, S., Bae, H. J., Barton, D. A., Beckett, L. A., Bell, J. M., Berman, S. E., Biessels, G. J., Black, S. E., Bos, I., Bowman, G. L., Brai, E., Brickman, A. M., Callahan, B. L., Corriveau, R. A., Fossati, S., Gottesman, R. F., Gustafson, D. R., Hachinski, V., … Hainsworth, A. H. (2019). White matter hyperintensities in vascular contributions to cognitive impairment and dementia (VCID): Knowledge gaps and opportunities. In Alzheimer’s and Dementia: Translational Research and Clinical Interventions (Vol. 5, pp. 107–117). 10.1016/j.trci.2019.02.001

Althubaity, N., Schubert, J., Martins, D., Yousaf, T., Nettis, M. A., Mondelli, V., Pariante, C., Harrison, N. A., Bullmore, E. T., Dima, D., Turkheimer, F. E., & Veronese, M. (2022). Choroid plexus enlargement is associated with neuroinflammation and reduction of blood brain barrier permeability in depression. NeuroImage: Clinical, 33. 10.1016/j.nicl.2021.102926

Aso, M., Kurachi, M., Suzuki, M., Yuasa, S., Matsui, M., & Saitoh, O. (1995). Asymmetry of the ventricle and age at the onset of schizophrenia. European Archives of Psychiatry and Clinical Neuroscience, 245(3), 142–144. 10.1007/BF02193086

Baker, C. M., Burks, J. D., Briggs, R. G., Conner, A. K., Glenn, C. A., Robbins, J. M., Sheets, J. R., Sali, G., McCoy, T. M., Battiste, J. D., O’Donoghue, D. L., & Sughrue, M. E. (2018). A Connectomic Atlas of the Human Cerebrum—Chapter 5: The Insula and Opercular Cortex. Operative Neurosurgery, 15(Suppl 1), S175–S244. 10.1093/ONS/OPY259

Baranger, D. A. A., Halchenko, Y. O., Satz, S., Ragozzino, R., Iyengar, S., Swartz, H. A., & Manelis, A. (2021a). Aberrant levels of cortical myelin distinguish individuals with depressive disorders from healthy controls. NeuroImage: Clinical, 32, 102790. 10.1016/j.nicl.2021.102790

Baranger, D. A. A., Halchenko, Y. O., Satz, S., Ragozzino, R., Iyengar, S., Swartz, H. A., & Manelis, A. (2021b). Protocol for a machine learning algorithm predicting depressive disorders using the T1w/T2w ratio. MethodsX, 8, 101595. 10.1016/j.mex.2021.101595

Benedetti, F., Aggio, V., Pratesi, M. L., Greco, G., & Furlan, R. (2020). Neuroinflammation in Bipolar Depression. In Frontiers in Psychiatry (Vol. 11). Frontiers Media S.A. 10.3389/fpsyt.2020.00071

Bourne, V. J. (2010). How are emotions lateralised in the brain? Contrasting existing hypotheses using the Chimeric Faces Test. Cognition and Emotion, 24(5), 903–911. 10.1080/02699930903007714

Brambilla, P., Harenski, K., Nicoletti, M., Mallinger, A. G., Frank, E., Kupfer, D. J., Keshavan, M. S., & Soares, J. C. (2001). MRI study of posterior fossa structures and brain ventricles in bipolar patients. Journal of Psychiatric Research, 35(6), 313–322. 10.1016/S0022-3956(01)00036-X

Chung, Y., Haut, K. M., He, G., van Erp, T. G. M., McEwen, S., Addington, J., Bearden, C. E., Cadenhead, K., Cornblatt, B., Mathalon, D. H., McGlashan, T., Perkins, D., Seidman, L. J., Tsuang, M., Walker, E., Woods, S. W., & Cannon, T. D. (2017). Ventricular enlargement and progressive reduction of cortical gray matter are linked in prodromal youth who develop psychosis. Schizophrenia Research, 189, 169–174. 10.1016/j.schres.2017.02.014

Coutu, J. P., Goldblatt, A., Rosas, H. D., & Salat, D. H. (2015). White matter changes are associated with ventricular expansion in aging, mild cognitive impairment, and Alzheimer’s disease. Journal of Alzheimer’s Disease, 49(2), 329–342. 10.3233/JAD-150306

Dale, A. M., Fischl, B., & Sereno, M. I. (1999). Cortical surface-based analysis. I. Segmentation and surface reconstruction. NeuroImage, 9(2), 179–194. 10.1006/nimg.1998.0395

Dantzer, R., O’Connor, J. C., Freund, G. G., Johnson, R. W., & Kelley, K. W. (2008). From inflammation to sickness and depression: When the immune system subjugates the brain. In Nature Reviews Neuroscience (Vol. 9, Issue 1). 10.1038/nrn2297

Davidson, R. J. (1993). Cerebral asymmetry and emotion: Conceptual and methodological conundrums. Cognition and Emotion, 7(1), 115–138. 10.1080/02699939308409180

Davidson, R. J., & Irwin, W. (1999). The functional neuroanatomy of emotion and affective style. In Trends in Cognitive Sciences (Vol. 3, Issue 1, pp. 11–21).Elsevier Current Trends. 10.1016/S1364-6613(98)01265-0

De Kovel, C. G. F., Aftanas, L., Aleman, A., Alexander-Bloch, A. F., Baune, B. T., Brack, I., Bülow, R., Filho, G. B., Carballedo, A., Connolly, C. G., Cullen, K. R., Dannlowski, U., Davey, C. G., Dima, D., Dohm, K., Erwin-Grabner, T., Frodl, T., Fu, C. H. Y., Hall, G. B., … Francks, C. (2019). No alterations of brain structural asymmetry in major depressive disorder: An ENIGMA consortium analysis. American Journal of Psychiatry, 176(12), 1039–1049. 10.1176/appi.ajp.2019.18101144

Dell’Osso, L., Armani, A., Rucci, P., Frank, E., Fagiolini, A., Corretti, G., Shear, M. K., Grochocinski, V. J., Maser, J. D., Endicott, J., & Cassano, G. B. (2002). Measuring mood spectrum: comparison of interview (SCI-MOODS) and self-report (MOODS-SR) instruments. Comprehensive Psychiatry, 43(1), 69–73.

Devorak, J., Torres-Platas, S. G., Davoli, M. A., Prud’homme, J., Turecki, G., & Mechawar, N. (2015). Cellular and molecular inflammatory profile of the choroid plexus in depression and suicide. Frontiers in Psychiatry, 6(OCT). 10.3389/fpsyt.2015.00138

Dilsaver, S. C. (2011). An estimate of the minimum economic burden of bipolar I and II disorders in the United States: 2009. Journal of Affective Disorders, 129(1–3), 79–83. 10.1016/j.jad.2010.08.030

Edmiston, E. E., Wang, F., Kalmar, J. H., Womer, F. Y., Chepenik, L. G., Pittman, B., Gueorguieva, R., Hur, E., Spencer, L., Staib, L. H., Constable, R. T., Fulbright, R. K., Papademetris, X., & Blumberg, H. P. (2011). Lateral ventricle volume and psychotic features in adolescents and adults with bipolar disorder. Psychiatry Research - Neuroimaging, 194(3). 10.1016/j.pscychresns.2011.07.005

Esteban, O., Birman, D., Schaer, M., Koyejo, O. O., Poldrack, R. A., & Gorgolewski, K. J. (2017). MRIQC: Advancing the automatic prediction of image quality in MRI from unseen sites. PloS One. 10.1371/journal.pone.0184661

Felger, J. C., & Miller, A. H. (2020). Identifying Immunophenotypes of Inflammation in Depression: Dismantling the Monolith. In Biological Psychiatry (Vol. 88, Issue 2, pp. 136–138). 10.1016/j.biopsych.2020.04.024

First, M. B. (2015). Structured Clinical Interview for the DSM (SCID). In The Encyclopedia of Clinical Psychology. 10.1002/9781118625392.wbecp351

Fleischer, V., Gonzalez-Escamilla, G., Ciolac, D., Albrecht, P., Küry, P., Gruchot, J., Dietrich, M., Hecker, C., Müntefering, T., Bock, S., Oshaghi, M., Radetz, A., Cerina, M., Krämer, J., Wachsmuth, L., Faber, C., Lassmann, H., Ruck, T., Meuth, S. G., … Groppa, S. (2021). Translational value of choroid plexus imaging for tracking neuroinflammation in mice and humans. Proceedings of the National Academy of Sciences of the United States of America, 118(36). 10.1073/pnas.2025000118

Friedman, J., Hastie, T., & Tibshirani, R. (2010). Regularization paths for generalized linear models via coordinate descent. Journal of Statistical Software, 33(1), 1–22. 10.18637/jss.v033.i01

Glaser, R., & Kiecolt-Glaser, J. K. (2005). Stress-induced immune dysfunction: Implications for health. In Nature Reviews Immunology (Vol. 5, Issue 3, pp. 243–251). 10.1038/nri1571

Glasser, M. F., Coalson, T. S., Robinson, E. C., Hacker, C. D., Harwell, J., Yacoub, E., Ugurbil, K., Andersson, J., Beckmann, C. F., Jenkinson, M., Smith, S. M., & Van Essen, D. C. (2016). A multi-modal parcellation of human cerebral cortex. Nature, 536(7615), 171–178. 10.1038/nature18933

Glasser, M. F., Sotiropoulos, S. N., Wilson, J. A., Coalson, T. S., Fischl, B., Andersson, J. L., Xu, J., Jbabdi, S., Webster, M., Polimeni, J. R., Van Essen, D. C., & Jenkinson, M. (2013). The minimal preprocessing pipelines for the Human Connectome Project. NeuroImage, 80, 105–124. 10.1016/j.neuroimage.2013.04.127

Glasser, M. F., & van Essen, D. C. (2011). Mapping human cortical areas in vivo based on myelin content as revealed by T1- and T2-weighted MRI. Journal of Neuroscience, 31(32), 11597–11616. 10.1523/JNEUROSCI.2180-11.2011

Gray, J. P., Müller, V. I., Eickhoff, S. B., & Fox, P. T. (2020). Multimodal abnormalities of brain structure and function in major depressive disorder: A meta-analysis of neuroimaging studies. American Journal of Psychiatry, 177(5), 422–434. 10.1176/appi.ajp.2019.19050560

Greenberg, P. E., Fournier, A. A., Sisitsky, T., Simes, M., Berman, R., Koenigsberg, S. H., & Kessler, R. C. (2021). The Economic Burden of Adults with Major Depressive Disorder in the United States (2010 and 2018). PharmacoEconomics, 39(6), 653–665. 10.1007/s40273-021-01019-4

Grewal, S., McKinlay, S., Kapczinski, F., Pfaffenseller, B., & Wollenhaupt-Aguiar, B. (2023). Biomarkers of neuroprogression and late staging in bipolar disorder: A systematic review. In Australian and New Zealand Journal of Psychiatry (Vol. 57, Issue 3). 10.1177/00048674221091731

Halchenko, Y., Goncalves, M., Castello, M. V. di O., Ghosh, S., Hanke, M., Dae Amlien, I., Brett, M., Salo, T., Gorgolewski, C., pvelasco Stadler, J., Kaczmarzyk, J., Lee, J., Lurie, D., Pellman, J., Melo, B., Poldrack, B., Nielson, D., … Feingold, F. (2019). nipy/heudiconv: v0.5.4 [0.5.4] - 2019-04-29. 10.5281/ZENODO.2653784.

Hamilton, M. (1960). A rating scale for depression. Journal of Neurology, Neurosurgery, and Psychiatry, 23, 56–62. 10.1136/jnnp.23.1.56

Hassel, S., Almeida, J. R., Kerr, N., Nau, S., Ladouceur, C. D., Fissell, K., Kupfer, D. J., & Phillips, M. L. (2008). Elevated striatal and decreased dorsolateral prefrontal cortical activity in response to emotional stimuli in euthymic bipolar disorder: no associations with psychotropic medication load. Bipolar Disorders, 10(8), 916–927. 10.1111/j.1399-5618.2008.00641.x

Hauser, P., Matochik, J., Altshuler, L. L., Denicoff, K. D., Conrad, A., Li, X., & Post, R. M. (2000). MRI-based measurements of temporal lobe and ventricular structures in patients with bipolar I and bipolar II disorders. Journal of Affective Disorders, 60(1). 10.1016/S0165-0327(99)00154-8

Hibar, D. P., Westlye, L. T., Doan, N. T., Jahanshad, N., Cheung, J. W., Ching, C. R. K., Versace, A., Bilderbeck, A. C., Uhlmann, A., Mwangi, B., Krämer, B., Overs, B., Hartberg, C. B., Abe, C., Dima, D., Grotegerd, D., Sprooten, E., Ben, E., Jimenez, E., … Andreassen, O. A. (2018). Cortical abnormalities in bipolar disorder: An MRI analysis of 6503 individuals from the ENIGMA Bipolar Disorder Working Group. Molecular Psychiatry, 23(4), 932–942. 10.1038/mp.2017.73

Hibar, D. P., Westlye, L. T., Van Erp, T. G. M., Rasmussen, J., Leonardo, C. D., Faskowitz, J., Haukvik, U. K., Hartberg, C. B., Doan, N. T., Agartz, I., Dale, A. M., Gruber, O., Krämer, B., Trost, S., Liberg, B., Abé, C., Ekman, C. J., Ingvar, M., Landén, M., … Andreassen, O. A. (2016). Subcortical volumetric abnormalities in bipolar disorder. Molecular Psychiatry. 10.1038/mp.2015.227

Ho, T. C., Gutman, B., Pozzi, E., Grabe, H. J., Hosten, N., Wittfeld, K., Völzke, H., Baune, B., Dannlowski, U., Förster, K., Grotegerd, D., Redlich, R., Jansen, A., Kircher, T., Krug, A., Meinert, S., Nenadic, I., Opel, N., Dinga, R., … Schmaal, L. (2022). Subcortical shape alterations in major depressive disorder: Findings from the ENIGMA major depressive disorder working group. Human Brain Mapping, 43(1), 341–351. 10.1002/hbm.24988

Joormann, J., & Gotlib, I. H. (2010). Emotion regulation in depression: Relation to cognitive inhibition. Cognition and Emotion, 24(2), 281–298. 10.1080/02699930903407948

Kang, K., Kwak, K., Yoon, U., & Lee, J. M. (2018). Lateral Ventricle Enlargement and Cortical Thinning in Idiopathic Normal-pressure Hydrocephalus Patients. Scientific Reports, 8(1), 1–9. 10.1038/s41598-018-31399-1

Kempton, M. J., Salvador, Z., Munafò, M. R., Geddes, J. R., Simmons, A., Frangou, S., & Williams, S. C. R. (2011). Structural neuroimaging studies in major depressive disorder: Meta-analysis and comparison with bipolar disorder. Archives of General Psychiatry, 68(7), 675–690. 10.1001/archgenpsychiatry.2011.60

Kieling, C., Buchweitz, C., Caye, A., Silvani, J., Ameis, S. H., Brunoni, A. R., Cost, K. T., Courtney, D. B., Georgiades, K., Merikangas, K. R., Henderson, J. L., Polanczyk, G. V., Rohde, L. A., Salum, G. A., & Szatmari, P. (2024). Worldwide Prevalence and Disability From Mental Disorders Across Childhood and Adolescence Evidence From the Global Burden of Disease Study. JAMA Psychiatry. 10.1001/jamapsychiatry.2023.5051

Klistorner, S., Barnett, M. H., Parratt, J., Yiannikas, C., Graham, S. L., & Klistorner, A. (2022). Choroid plexus volume in multiple sclerosis predicts expansion of chronic lesions and brain atrophy. Annals of Clinical and Translational Neurology, 9(10). 10.1002/acn3.51644

Li, X., Morgan, P. S., Ashburner, J., Smith, J., & Rorden, C. (2016). The first step for neuroimaging data analysis: DICOM to NIfTI conversion. Journal of Neuroscience Methods, 264, 47–56. 10.1016/j.jneumeth.2016.03.001

Lizano, P., Lutz, O., Ling, G., Lee, A. M., Eum, S., Bishop, J. R., Kelly, S., Pasternak, O., Clementz, B., Pearlson, G., Sweeney, J. A., Gershon, E., Tamminga, C., & Keshavan, M. (2019). Association of choroid plexus enlargement with cognitive, inflammatory, and structural phenotypes across the psychosis spectrum. American Journal of Psychiatry, 176(7). 10.1176/appi.ajp.2019.18070825

Maller, J. J., Anderson, R., Thomson, R. H., Rosenfeld, J. V., Daskalakis, Z. J., & Fitzgerald, P. B. (2015). Occipital bending (Yakovlevian torque) in bipolar depression. Psychiatry Research - Neuroimaging, 231(1), 8–14. 10.1016/j.pscychresns.2014.11.008

Maller, J. J., Thomson, R. H. S., Rosenfeld, J. V., Anderson, R., Daskalakis, Z. J., & Fitzgerald, P. B. (2014). Occipital bending in depression. Brain, 137(6), 1830–1837. 10.1093/brain/awu072

Manelis, A., Almeida, J. R. C., Stiffler, R., Lockovich, J. C., Aslam, H. A., & Phillips, M. L. (2016). Anticipation-related brain connectivity in bipolar and unipolar depression: A graph theory approach. Brain, 139(9), 2554–2566. 10.1093/brain/aww157

Manelis, A., Iyengar, S., Swartz, H. A., & Phillips, M. L. (2020). Prefrontal cortical activation during working memory task anticipation contributes to discrimination between bipolar and unipolar depression. Neuropsychopharmacology, 45(6), 956–963. 10.1038/s41386-020-0638-7

Mierzwa, A. J., Marion, C. M., Sullivan, G. M., McDaniel, D. P., & Armstrong, R. C. (2015). Components of myelin damage and repair in the progression of white matter pathology after mild traumatic brain injury. Journal of Neuropathology and Experimental Neurology, 74(3). 10.1097/NEN.0000000000000165

Miola, A., Cattarinussi, G., Antiga, G., Caiolo, S., Solmi, M., & Sambataro, F. (2022). Difficulties in emotion regulation in bipolar disorder: A systematic review and meta-analysis. Journal of Affective Disorders, 302, 352–360. 10.1016/j.jad.2022.01.102

Murck, H., Fava, M., Cusin, C., Fatt, C. C., & Trivedi, M. (2024). Brain ventricle and choroid plexus morphology as predictor of treatment response in major depression: Findings from the EMBARC study. Brain, Behavior, and Immunity - Health, 35. 10.1016/j.bbih.2023.100717

Murck, H., Lehr, L., & Jezova, D. (2023). A viewpoint on aldosterone and BMI related brain morphology in relation to treatment outcome in patients with major depression. In Journal of Neuroendocrinology (Vol. 35, Issue 2). 10.1111/jne.13219

Nelson, H. E. (1982). National Adult Reading Test (NART): For the Assessment of Premorbid Intelligence in Patients with Dementia: Test Manual. In 1982. Nfer-Nelson.

Okada, N., Fukunaga, M., Miura, K., Nemoto, K., Matsumoto, J., Hashimoto, N., Kiyota, M., Morita, K., Koshiyama, D., Ohi, K., Takahashi, T., Koeda, M., Yamamori, H., Fujimoto, M., Yasuda, Y., Hasegawa, N., Narita, H., Yokoyama, S., Mishima, R., … Hashimoto, R. (2023). Subcortical volumetric alterations in four major psychiatric disorders: a mega-analysis study of 5604 subjects and a volumetric data-driven approach for classification. Molecular Psychiatry. 10.1038/s41380-023-02141-9

Pereira, A. C., Oliveira, J., Silva, S., Madeira, N., Pereira, C. M. F., & Cruz, M. T. (2021). Inflammation in Bipolar Disorder (BD): Identification of new therapeutic targets. In Pharmacological Research (Vol. 163). 10.1016/j.phrs.2020.105325

Qiu, M., Liu, G., Zhang, H., Huang, Y., Ying, S., Wang, J., Shen, T., & Peng, D. (2020). The Insular Subregions Topological Characteristics of Patients With Bipolar Depressive Disorder. Frontiers in Psychiatry, 11. 10.3389/fpsyt.2020.00253

Ricigliano, V. A. G., Morena, E., Colombi, A., Tonietto, M., Hamzaoui, M., Poirion, E., Bottlaender, M., Gervais, P., Louapre, C., Bodini, B., & Stankoff, B. (2021). Choroid plexus enlargement in inflammatory multiple sclerosis: 3.0-T MRI and translocator protein PET evaluation. Radiology, 301(1). 10.1148/radiol.2021204426

Robinson, E. C., Garcia, K., Glasser, M. F., Chen, Z., Coalson, T. S., Makropoulos, A., Bozek, J., Wright, R., Schuh, A., Webster, M., Hutter, J., Price, A., Cordero Grande, L., Hughes, E., Tusor, N., Bayly, P. V., Van Essen, D. C., Smith, S. M., Edwards, A. D., … Rueckert, D. (2018). Multimodal surface matching with higher-order smoothness constraints. NeuroImage, 167, 453–465. 10.1016/j.neuroimage.2017.10.037

Rolls, E. T., Cheng, W., & Feng, J. (2020). The orbitofrontal cortex: Reward, emotion and depression. Brain Communications, 2(2). 10.1093/braincomms/fcaa196

Rosner, B. (1983). American Society for Quality Percentage Points for a Generalized ESD Many-Outlier Procedure. Source: Technometrics, 25(2).

Sacchet, M. D., & Gotlib, I. H. (2017). Myelination of the brain in major depressive disorder: An in vivo quantitative magnetic resonance imaging study. Scientific Reports, 7(1). 10.1038/s41598-017-02062-y

Scelsi, C. L., Rahim, T. A., Morris, J. A., Kramer, G. J., Gilbert, B. C., & Forseen, S. E. (2020). The lateral ventricles: A detailed review of anatomy, development, and anatomic variations. American Journal of Neuroradiology, 41(4), 566–572. 10.3174/AJNR.A6456

Schmaal, L., Veltman, D. J., Van Erp, T. G. M., Smann, P. G., Frodl, T., Jahanshad, N., Loehrer, E., Tiemeier, H., Hofman, A., Niessen, W. J., Vernooij, M. W., Ikram, M. A., Wittfeld, K., Grabe, H. J., Block, A., Hegenscheid, K., Völzke, H., Hoehn, D., Czisch, M., … Hibar, D. P. (2016). Subcortical brain alterations in major depressive disorder: Findings from the ENIGMA Major Depressive Disorder working group. Molecular Psychiatry, 21(6). 10.1038/mp.2015.69

Scott, M. L., Golden, C. J., Ruedrich, S. L., & Bishop, R. J. (1983). Ventricular enlargement in major depression. Psychiatry Research, 8(2). 10.1016/0165-1781(83)90095-1

Seghier, M. L. (2008). Laterality index in functional MRI: methodological issues. Magnetic Resonance Imaging, 26(5), 594–601. 10.1016/j.mri.2007.10.010

Shim, J. M., Cho, S. E., Kang, C. K., & Kang, S. G. (2023). Low myelin-related values in the fornix and thalamus of 7 Tesla MRI of major depressive disorder patients. Frontiers in Molecular Neuroscience, 16. 10.3389/fnmol.2023.1214738

Simons, M., & Nave, K. A. (2016). Oligodendrocytes: Myelination and axonal support. In Cold Spring Harbor Perspectives in Biology (Vol. 8, Issue 1, pp. 1–16). 10.1101/cshperspect.a020479

Sinnecker, T., Ruberte, E., Schädelin, S., Canova, V., Amann, M., Naegelin, Y., Penner, I. K., Müller, J., Kuhle, J., Décard, B., Derfuss, T., Kappos, L., Granziera, C., Wuerfel, J., Magon, S., & Yaldizli, Ö. (2020). New and enlarging white matter lesions adjacent to the ventricle system and thalamic atrophy are independently associated with lateral ventricular enlargement in multiple sclerosis. Journal of Neurology, 267(1). 10.1007/s00415-019-09565-w

Slavich, G. M., & Irwin, M. R. (2014). From stress to inflammation and major depressive disorder: A social signal transduction theory of depression. Psychological Bulletin, 140(3), 774–815. 10.1037/a0035302

Strakowski, S. M., DelBello, M. P., Zimmerman, M. E., Getz, G. E., Mills, N. P., Ret, J., Shear, P., & Adler, C. M. (2002). Ventricular and periventricular structural volumes in first-versus multiple-episode bipolar disorder. American Journal of Psychiatry, 159(11), 1841–1847. 10.1176/appi.ajp.159.11.1841

Tadayon, E., Pascual-Leone, A., Press, D., & Santarnecchi, E. (2020). Choroid plexus volume is associated with levels of CSF proteins: relevance for Alzheimer’s and Parkinson’s disease. Neurobiology of Aging, 89. 10.1016/j.neurobiolaging.2020.01.005

Thompson, D., Brissette, C. A., & Watt, J. A. (2022). The choroid plexus and its role in the pathogenesis of neurological infections. In Fluids and Barriers of the CNS (Vol. 19, Issue 1). BMC. 10.1186/s12987-022-00372-6

Tonietto, M., Poirion, E., Lazzarotto, A., Ricigliano, V., Papeix, C., Bottlaender, M., Bodini, B., & Stankoff, B. (2023). Periventricular remyelination failure in multiple sclerosis: a substrate for neurodegeneration. Brain, 146(1), 182–194. 10.1093/brain/awac334

Troubat, R., Barone, P., Leman, S., Desmidt, T., Cressant, A., Atanasova, B., Brizard, B., El Hage, W., Surget, A., Belzung, C., & Camus, V. (2021). Neuroinflammation and depression: A review. European Journal of Neuroscience, 53(1), 151–171. 10.1111/ejn.14720

Turkheimer, F. E., Althubaity, N., Schubert, J., Nettis, M. A., Cousins, O., Dima, D., Mondelli, V., Bullmore, E. T., Pariante, C., & Veronese, M. (2021). Increased serum peripheral C-reactive protein is associated with reduced brain barriers permeability of TSPO radioligands in healthy volunteers and depressed patients: implications for inflammation and depression. Brain, Behavior, and Immunity, 91, 487–497. 10.1016/j.bbi.2020.10.025

Turnbull, O. H., & Salas, C. E. (2021). The neuropsychology of emotion and emotion regulation: The role of laterality and hierarchy. In Brain Sciences (Vol. 11, Issue 8, p. 1075). Multidisciplinary Digital Publishing Institute. 10.3390/brainsci11081075

Turner, C. A., Thompson, R. C., Bunney, W. E., Schatzberg, A. F., Barchas, J. D., Myers, R. M., Akil, H., & Watson, S. J. (2014). Altered choroid plexus gene expression in major depressive disorder. Frontiers in Human Neuroscience, 8(1 APR). 10.3389/fnhum.2014.00238

Valdés-Tovar, M., Rodríguez-Ramírez, A. M., Rodríguez-Cárdenas, L., Sotelo-Ramírez, C. E., Camarena, B., Sanabrais-Jiménez, M. A., Solís-Chagoyán, H., Argueta, J., & López-Riquelme, G. O. (2022). Insights into myelin dysfunction in schizophrenia and bipolar disorder. World Journal of Psychiatry, 12(2), 264–285. 10.5498/wjp.v12.i2.264

Visconti di Oleggio Castello, M., Dobson, J. E., Sackett, T., Kodiweera, C., Haxby, J. V., Goncalves, M., Ghosh, S., & Halchenko, Y. O. (2020). ReproNim/reproin 0.6.0. 10.5281/ZENODO.3625000

Xie, C., Jia, T., Rolls, E. T., Robbins, T., Sahakian, B. J., Zhang, J., Liu, Z., Cheng, W., Luo, Q., Zac Lo, C. Y., Wang, H., Banaschewski, T., Barker, G. J., Bokde, A. L. W., Büchel, C., Quinlan, E. B., Desrivières, S., Flor, H., Grigis, A., … Zhang, Y. (2021). Reward Versus Nonreward Sensitivity of the Medial Versus Lateral Orbitofrontal Cortex Relates to the Severity of Depressive Symptoms. Biological Psychiatry: Cognitive Neuroscience and Neuroimaging, 6(3), 259–269. 10.1016/j.bpsc.2020.08.017

Yamada, S., & Mase, M. (2023). Cerebrospinal Fluid Production and Absorption and Ventricular Enlargement Mechanisms in Hydrocephalus. Neurologia Medico-Chirurgica, 63(4), 141–151. 10.2176/jns-nmc.2022-0331

Young, R. C., Biggs, J. T., Ziegler, V. E., & Meyer, D. A. (1978). A rating scale for mania: Reliability, validity and sensitivity. British Journal of Psychiatry, 133(11), 429–435. 10.1192/bjp.133.5.429

Zhang, T. J., Wu, Q. Z., Huang, X. Q., Sun, X. L., Zou, K., Lui, S., Liu, F., Hu, J. M., Kuang, W. H., Li, D. M., Li, F., Chen, H. F., Chan, R. C. K., Mechelli, A., & Gong, Q. Y. (2009). Magnetization transfer imaging reveals the brain deficit in patients with treatment-refractory depression. Journal of Affective Disorders, 117(3), 157–161. 10.1016/j.jad.2009.01.003

Zou, H., & Hastie, T. (2005). Regularization and variable selection via the elastic net. Journal of the Royal Statistical Society. Series B: Statistical Methodology, 67(2), 301–320. 10.1111/j.1467-9868.2005.00503.x

Zuo, Z., Ran, S., Wang, Y., Li, C., Han, Q., Tang, Q., Qu, W., & Li, H. (2019). Asymmetry in cortical thickness and subcortical volume in treatment-naïve major depressive disorder. NeuroImage: Clinical, 21, 101614. 10.1016/j.nicl.2018.101614

